# Identification, analysis and prediction of valid and false information related to vaccines from Romanian tweets

**DOI:** 10.1101/2023.08.19.23294319

**Authors:** Andrei Valeanu, Dragos Paul Mihai, Corina Andrei, Ciprian Puscasu, Alexandra Mihaela Ionica, Miruna Ioana Hinoveanu, Valentina Patricia Predoi, Ema Bulancea, Cornel Chirita, Simona Negres, Cristian Daniel Marineci

**Author notes:** Corresponding author: (DPM).

## Abstract

The online misinformation might undermine the vaccination efforts. Therefore, given the fact that no study specifically analyzed online vaccine related content written in Romanian, the main objective of the study was to detect and evaluate tweets related to vaccines and written in Romanian language. 1400 Romanian vaccine related tweets were manually classified in true, neutral and fake information and analyzed based on wordcloud representations, a correlation analysis between the three classes and specific tweet characteristics and the validation of several predictive machine learning algorithms. The tweets annotated as misinformation showed specific word patterns and were liked and reshared more often as compared to the true and neutral ones. The validation of the machine learning algorithms yielded enhanced results in terms of Area Under the Receiver Operating Characteristic Curve Score (0.744-0.843) when evaluating the Support Vector Classifier. The predictive model estimates in a well calibrated manner the probability that a specific Twitter post is true, neutral or fake. The current study offers important insights regarding vaccine related online content written in an Eastern European language. Future studies must aim at building an online platform for rapid identification of vaccine misinformation and raising awareness for the general population.

## 1. Introduction

Vaccines are among the most important medications worldwide. It is estimated that they have saved millions of lives and that they will continue to do so [1]. Vaccines had a crucial role in the eradication of smallpox in 1980 and in bringing poliomyelitis very close to eradication [2,3]. In addition, a report found that as of November 2021, the Covid-19 vaccines saved nearly half a million lives in less than a year in the over 60 years old group across the WHO European Region [4].

However, despite their essential therapeutic effect and good safety profile, various disinformation articles, news and social media posts have emerged in the last decades, leading to the anti-vaccine movement. Even though the facts behind such information were proven to be false, the vaccine fake news phenomenon has led in many countries to a reduction of the vaccination rates, both in the adult and the pediatric population [5]. Low vaccination rates pose the risk of diseases that currently have a low impact in the population to return with a higher impact, with an additional burden on the healthcare system [1].

Numerous fake news related to the Covid-19 vaccines have also emerged and spread during the pandemic [5,6]. The online disinformation, in combination with other social and economic factors (such as media usage, educational background, health literacy, public trust in the government and health system) have been hypothesized to influence a person’s decision of getting the Covid-19 vaccine [7–9].

Social media platforms (such as Facebook and Twitter) are among the most important tools for spreading information about vaccines, whether it is valid information or fake news [5]. Therefore, the analysis of the content distributed through such platforms might be of an utmost importance in order to inform the general population and the health policy makers.

With regards to Twitter content, several studies have evaluated vaccine related posts (whether Covid or non-Covid), with regards to identifying and predicting disinformation, analyzing vaccine hesitancy, performing sentiment classification or other relevant analyses [10–19]. The majority of the studies were based on tweets written in English. Other analyzed languages were Dutch, Moroccan and Turkish, while one study involved a multi-language approach for detection and classification of tweets related to Covid-19 [12,14,15,18]. However, to our knowledge, no such study specifically analyzed vaccine related content based on Romanian tweets.

Therefore, the objective of the present study was to analyze vaccine related content, with the main goal of developing specific machine learning models for predicting disinformation from tweets written in Romanian.

## 2. Materials and Methods

### 2.1. Data collection

The vaccine related tweets were automatically extracted by using snscrape package developed in Python programming language [20,21]. The Twitter API was queried by using all the Romanian forms of the noun “vaccine”, the verb “to vaccinate”, as well as other ironical related terms (such as “vax”, “vaxin” or “vaxxin”) [20]. All tweets (both original posts and replies, both Covid and non-Covid vaccine information) from 4 relevant 4-week periods during the Covid-19 pandemic were initially collected (First period: March 16, 2020 – April 12, 2020; second period: December 27, 2020 – January 23, 2021; third period: May 3, 2021 – May 30, 2021; fourth period: October 18, 2021 – November 14, 2021). Each period was considered suggestive for the aim of extracting relevant batches of tweets. March 16, 2020 was the date in which the Emergency State was declared in Romania due to the Covid-19 pandemic; December 27, 2020 was the first day of the Covid-19 vaccination campaign in Romania; May 2021 was the month in which the highest average number of Covid-19 vaccine doses were administered and October-November 2021 was the period with the highest number of deaths due to Covid-19 in Romania [22].

After the initial collection, for each of the 4 periods, the tweets from the 7-day period with the highest number of tweets were considered and represented the internal dataset (1300 tweets). The final collection stage also included selecting the tweets with at least one retweet. The two filters were applied in order to obtain a relevant batch of posts related to vaccines, feasible for manual annotation [20,21]. In addition, a random batch of 100 tweets from April 2021 were collected, which represented the external validation dataset.

In order to obtain relevant information for the data analysis phase, the following parameters were extracted for each tweet: date and time, tweet ID, tweet content, number of likes, number of retweets, number of replies. All the information was anonymously collected through snscrape package, which is based on the Twitter API [20].

### 2.2. Manual annotation

In order to analyze the collected Twitter posts in a relevant manner, all the tweets had to be manually annotated. The tweets were classified in true, neutral or fake based on their text content. The true classification (class 0) meant valid scientific information related to vaccines (whether it was about Covid-19 vaccines or other vaccine types) or true general information regarding the Romanian vaccination campaign. The neutral classification (class 1) regarded irrelevant, ironical or other vaccine related comments, without manipulative or misleading intent. The fake classification (class 2) referred to false or misleading information related to vaccines (both Covid-19 and other types) or the Romanian vaccination campaign. The scientific validity of the posts, when appropriate, was assessed in relation to the official sources of health information (such as the European Centre for Disease Prevention and Control, the Summaries of Product Characteristics of the vaccines approved in Romania or trusted health fact check websites) [23–27]. It should also be noted that the majority of the tweets were related to Covid vaccines. However, the Twitter posts related to other types of vaccines were not eliminated, in order to increase the variability and complexity of the obtained dataset.

A total number of 9 annotators participated in the task. The external validation dataset was assessed by all 9 annotators and the final classification of each tweet was obtained by a majority vote. The internal dataset was annotated in a similar manner; however, due to the larger number of posts, the internal data was split into 3 parts of similar number of tweets and each part was annotated by 3 different annotators and the final classification was established by a majority vote. Hence, all 9 annotators took part both in the external validation data and in the internal data annotation. In addition, it should be mentioned that when a majority vote could not be applied (due to an equal distribution of votes among the three classes), the tweet was annotated as neutral, in order to ensure an unbiased data analysis. No tweet was eliminated when a majority vote could not be applied, in order to enhance the variability of the processed vaccine data. The agreement between annotators was established on the internal and external data by using Krippendorff’s alpha coefficient. The computation of the metric was considered relevant since it provides an ordinal option when assessing the agreement [28]. Therefore, the differences between true and neutral annotations are not penalized as hard as the differences between true and fake annotations.

### 2.3. Text preprocessing

In order to accurately analyze the annotated tweets, the text content had to be preprocessed. The text preprocessing and machine learning development and validation were performed by using Python Programming Language, version 3.9.2 [21].

In order to curate the text and obtain a simplified version, all special characters and stop words were removed from the tweets and all letters were converted to lowercase. The standard stop word list for Romanian provided by spacy was used. In addition, with the aim of providing a bias reduction for the development of the machine learning algorithm, all hyperlinks and words starting with the ‘@’ symbol (with which the content of tweet replies begins) were also eliminated. However, it should be noted that no lemmatization was performed on the selected tweets, since, taking into consideration practical reasons, it was considered that different word forms might provide different meaning and intent to specific phrases; moreover, as an example, as opposed to English language, the Romanian language has a higher number of forms for the noun “vaccine” and the verb “to vaccinate” [29,30].

### 2.4. Preliminary analysis

In order to characterize and extract relevant characteristics from the obtained dataset, a preliminary analysis was performed, based on two important methods. The first one implied extracting the most frequent single words and word combinations based on a wordcloud technique, in order to offer a simplified and relevant visualization of the dataset The words were obtained for each of the 3 classes (true, neutral and fake) from the 1300 vaccine tweets. The second method was applied in order to evaluate the relationships between the manual classification of the Twitter posts and other characteristics. Hence the Spearman’s correlation coefficient, along with the p value for statistical significance were computed between the manual classification (true – class 0, neutral – class 1, fake – class 2) and each of the following characteristics of the 1300 tweets: number of replies, number of retweets, number of likes and the length of the post, quantified by the number of words [31].

### 2.5. Building and validating the machine learning algorithm

The machine learning algorithm was developed by using Python’s scikit-learn package (four classical machine learning models: Support Vector Machines Classifier (SVM), Multilayer Perceptron (MLP, a type of neural networks), Random Forest Classifier (RFC) and an ensemble model (scikit-learn Voting Classifier), developed by averaging the probabilities which were predicted by the SVM and the MLP), as well as Tensorflow (for two specific deep learning models: recurrent convolutional neural networks (RCNN) – Tensorflow implementation and BERT – based on a model which was pretrained on a Romanian 15 GB uncased text corpus, downloaded from Huggingface (dumitrescustefan/bert-base-romanian-uncased-v1 model) and then executed through Tensorflow) [32–34]. With regards to the classical machine learning models since scikit-learn does not accept string data as input, the text content had to be converted to numerical data, by using the TfidfVectorizer function. No words were eliminated from the text corpus when performing the string-to-float conversion [34]. On the other hand, the deep learning models which were implemented required specific word tokenizers. The RCNN model was built after using the specific Tensorflow tokenizer, while the BERT model implemented the specific Romanian based AutoTokenizer downloaded from the huggingface website [32,33].

The six machine learning algorithms were validated and compared on the obtained data. They were tested based on their ability of estimating the probability that a specific tweet is true, neutral or fake, as well as of correctly classifying a tweet as being true, neutral or fake. The Area Under the Receiver Operating Characteristic Curve Score (ROC AUC Score, both a One-Versus-One (OVO) strategy and a One-Versus-Rest (OVR) strategy) was used for testing the probability prediction ability of the algorithms and was the most important overall measure for evaluating the machine learning models: the higher the ROC AUC Score is, the better are the probabilities calibrated. In addition, the accuracy, precision, recall, F1 Score and Matthews Correlation Coefficient were used to test the classification ability of the developed models. The Matthews Correlation Coefficient was considered the most important global classification measure, since it provides a relevant bias reduction approach and takes into consideration class imbalance [34–36].

Both an internal and an external validation were performed for the machine learning algorithms. The internal validation was performed on the 1300 tweets (internal dataset) and aimed at evaluating the internal consistency of the model combined with the ability of perform on unseen data. Hence, the dataset was split into 4 parts based on the 4 pandemic periods for which the posts were collected (internal period validation). The predictive algorithms were validated 4 times: each time, the training set included the tweets from 3 of the periods; the model was trained on the 3 periods and was evaluated based on the unseen data from the 4^th^ period. Therefore, the model was trained and validated until all the 4 periods represented in turn the test set. In addition, a repeated 5-fold cross-validation (with 10 iterations) was also performed [32–34]. However, the internal period validation strategy was considered much more relevant than the cross-validation, since all tweets from a specific period were either in the training set or the test set and the risk that the model was evaluated on similar tweets was significantly reduced.

The external validation of the algorithms implied training the models on the internal data and evaluating their performance on the external dataset represented by the 100 tweets from April 2021 [32–34].

Figure 1 briefly presents the three main strategies within the validation process of the machine learning algorithms.

**Fig 1.**
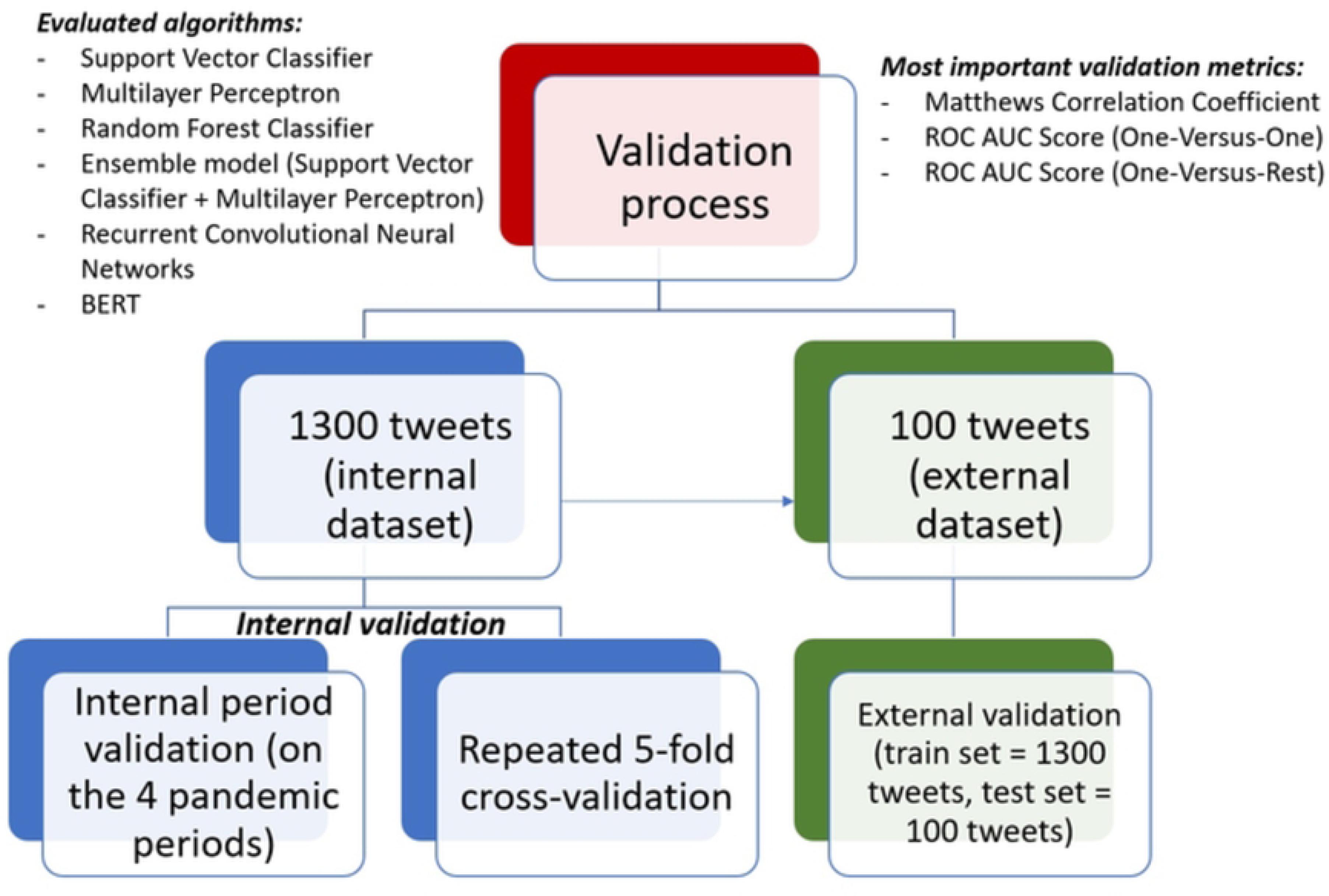
The validation process of the machine learning algorithms.

The final model (SVM) for future identification of specific vaccine tweets was chosen based on the best results obtained in terms of OVO and OVR ROC AUC Scores and was built by taking into consideration the internal dataset (1300 tweets). The model was implemented based on a probabilistic approach (useful for reliable probability estimation), a radial basis function (RBF) kernel, a penalty parameter of the error term (C value) set to 1, while reducing bias caused by class imbalance and breaking ties according to the confidence values of the RBF. In addition, a detailed analysis was undertaken based on the probability predictions of the final model on 3 tweets from the external data (one true post, one neutral post and one fake post) [34,37].

## 3. Results

### 3.1. Data collection

A total number of 1344 tweets were obtained, of which 44 were eliminated due to content in other languages. An additional 100 tweets were randomly selected from another period (April 2021, from the 7-day period with the highest number of tweets, independent on the number of retweets) and represented the dataset for external validation. Table 1 presents the final 7-day periods from which the posts were collected, as well as the number of tweets for each weekly time interval.

**Table 1.**
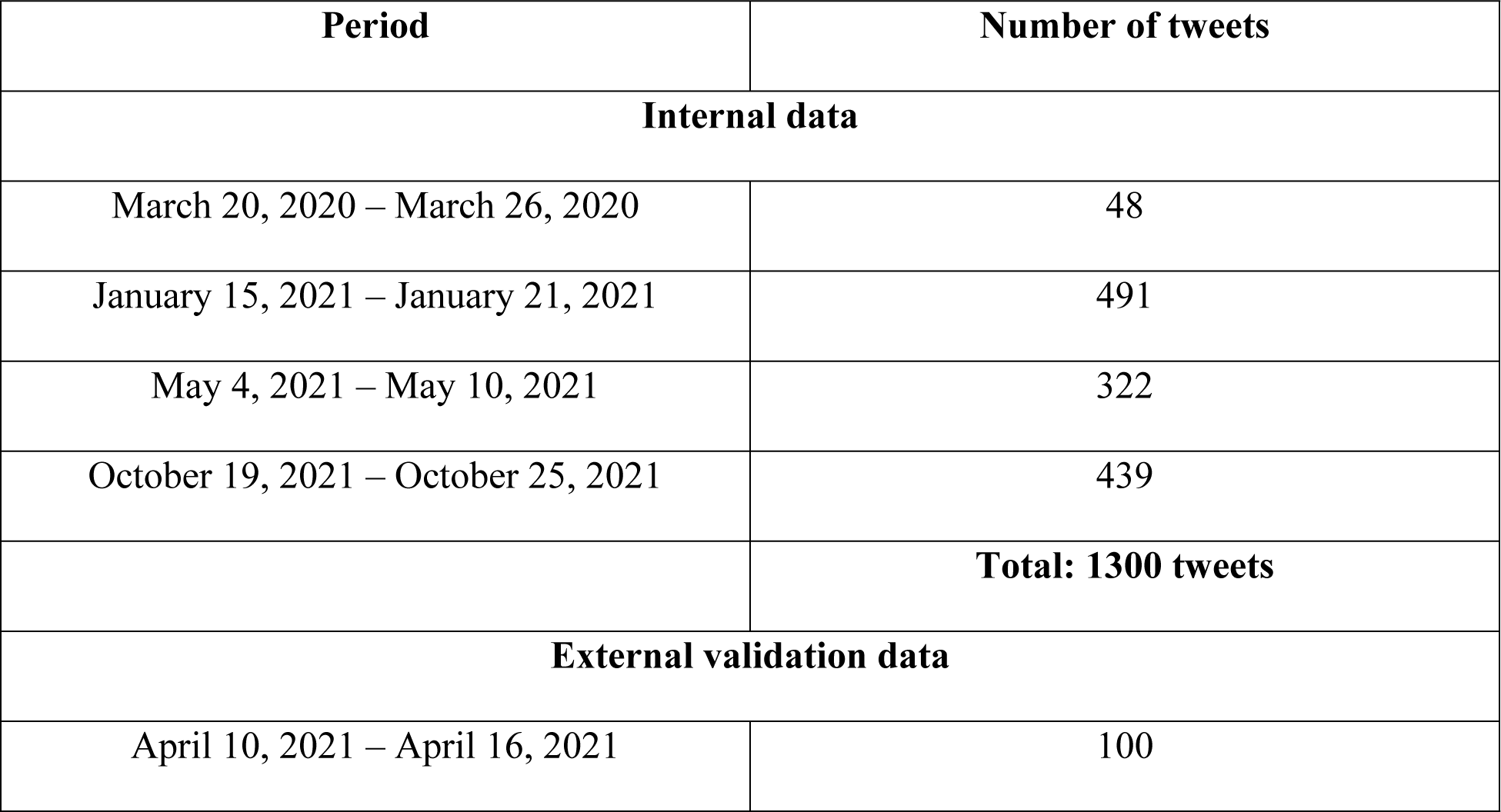
The time periods corresponding to the collected vaccine related tweets.

| Period | Number of tweets |
| --- | --- |
| **Internal data** |  |
| March 20, 2020 – March 26, 2020 | 48 |
| January 15, 2021 – January 21, 2021 | 491 |
| May 4, 2021 – May 10, 2021 | 322 |
| October 19, 2021 – October 25, 2021 | 439 |
|  | **Total: 1300 tweets** |
| **External validation data** |  |
| April 10, 2021 – April 16, 2021 | 100 |

### 3.2. Manual annotation

The manual annotation yielded an average inter-agreement Krippendorff’s alpha of 0.64 for the internal dataset (0.69 for Team 1, 0.58 for Team 2 and 0.64 for Team 3) and of 0.7 for the external dataset. After applying the majority vote rule, from the 1300 tweets (internal dataset), a total number of 488 (37.5%) were classified as true, 373 (28.7%) as neutral and 439 (33.8%) as fake. From the 100 tweets representing the external dataset, 53 (53%) were classified as true, 24 (24%) as neutral and 23 (23%) as fake.

In terms of overall inter-annotator agreement, from the 1300 tweets, 686 (52.8%) reached perfect agreement between the 3 annotators. From the 100 tweets from the external dataset, 15 (15%) reached perfect agreement between all 9 annotators.

### 3.3. Preliminary analysis

Table 2 presents the most relevant words and word combinations for the true, neutral and fake tweets within the internal dataset. The most relevant 7 words and word combinations (as considered by the annotators) of the most frequent 30 are presented. The words were translated from Romanian to English and the original Romanian version is also presented in parenthesis, when appropriate. Table 3 summarizes the results obtained by computing the Spearman’s correlation coefficient. The p values are not given, since all pairs yielded statistically significant results (p<0.05). Wordcloud representations of the most relevant words written in Romanian and graphical illustration of correlation analysis are shown in Figure 2.

**Fig 2.**
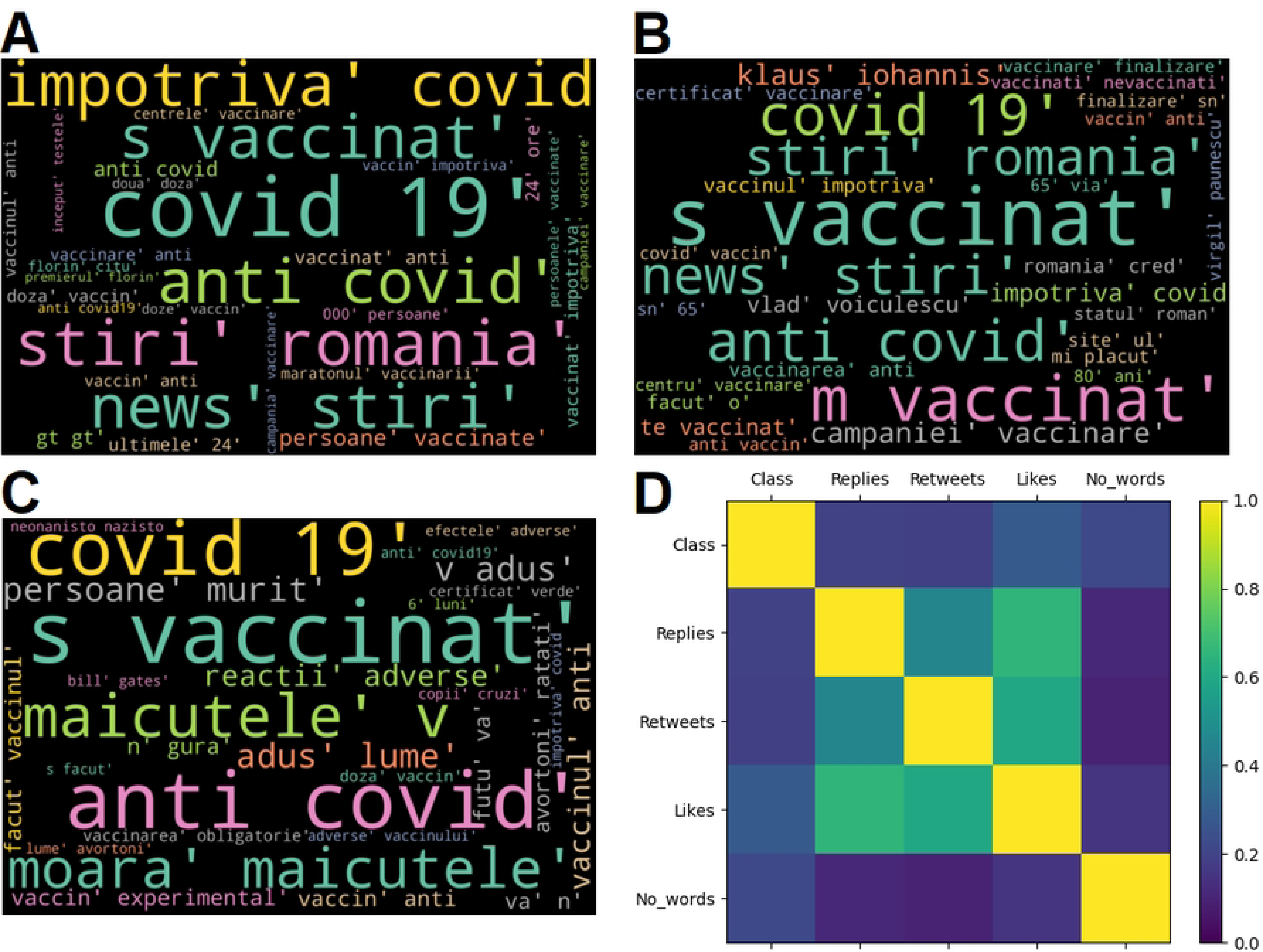
Wordcloud representation (the most relevant 30 words written in Romanian) for the tweets labelled as true (A), neutral (B) and fake (C); Correlation analysis results (D)

**Table 2.**
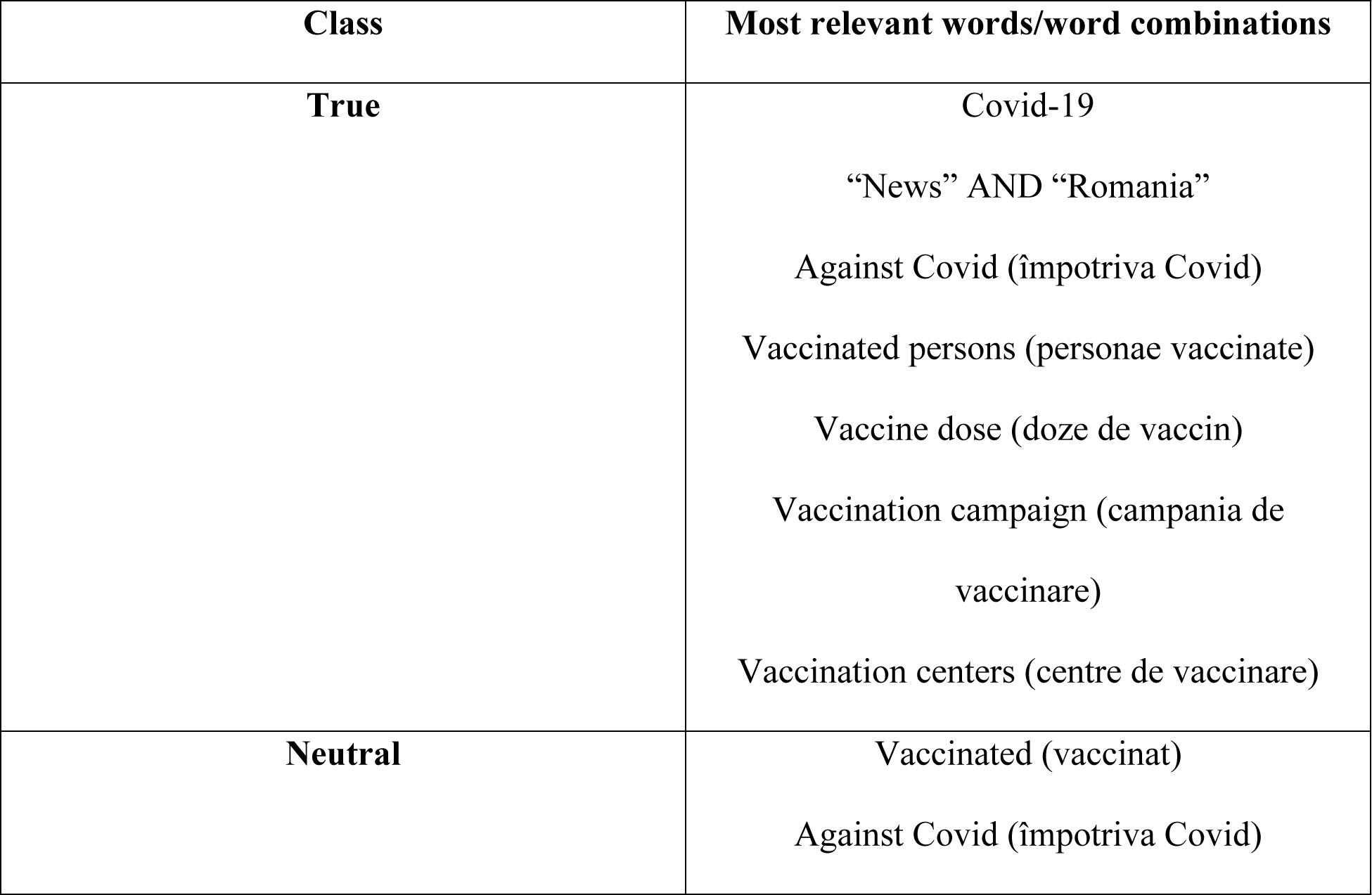
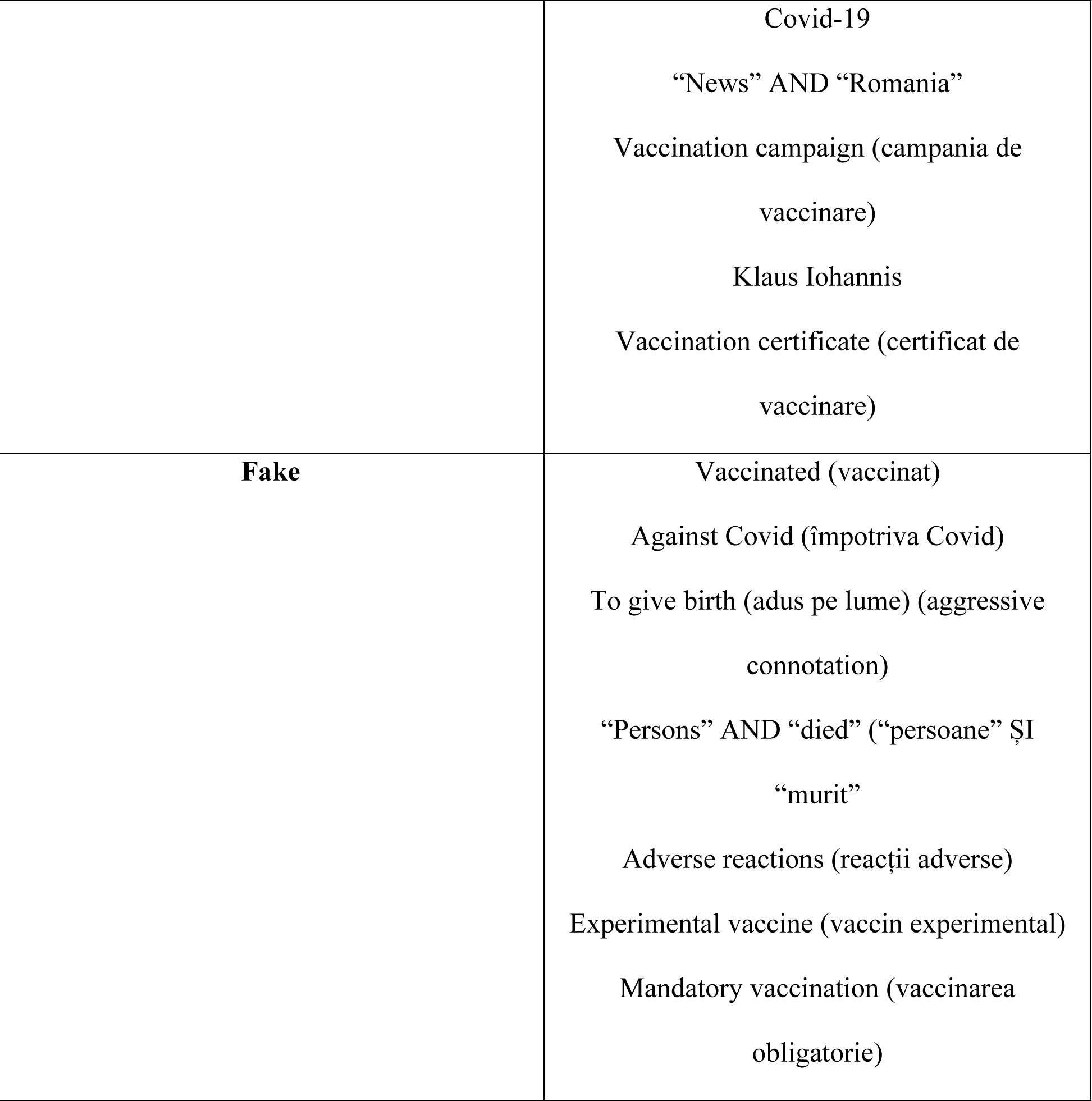
Most relevant words and word combinations identified through a wordcloud model for each of the 3 classes.

**Table 3.**
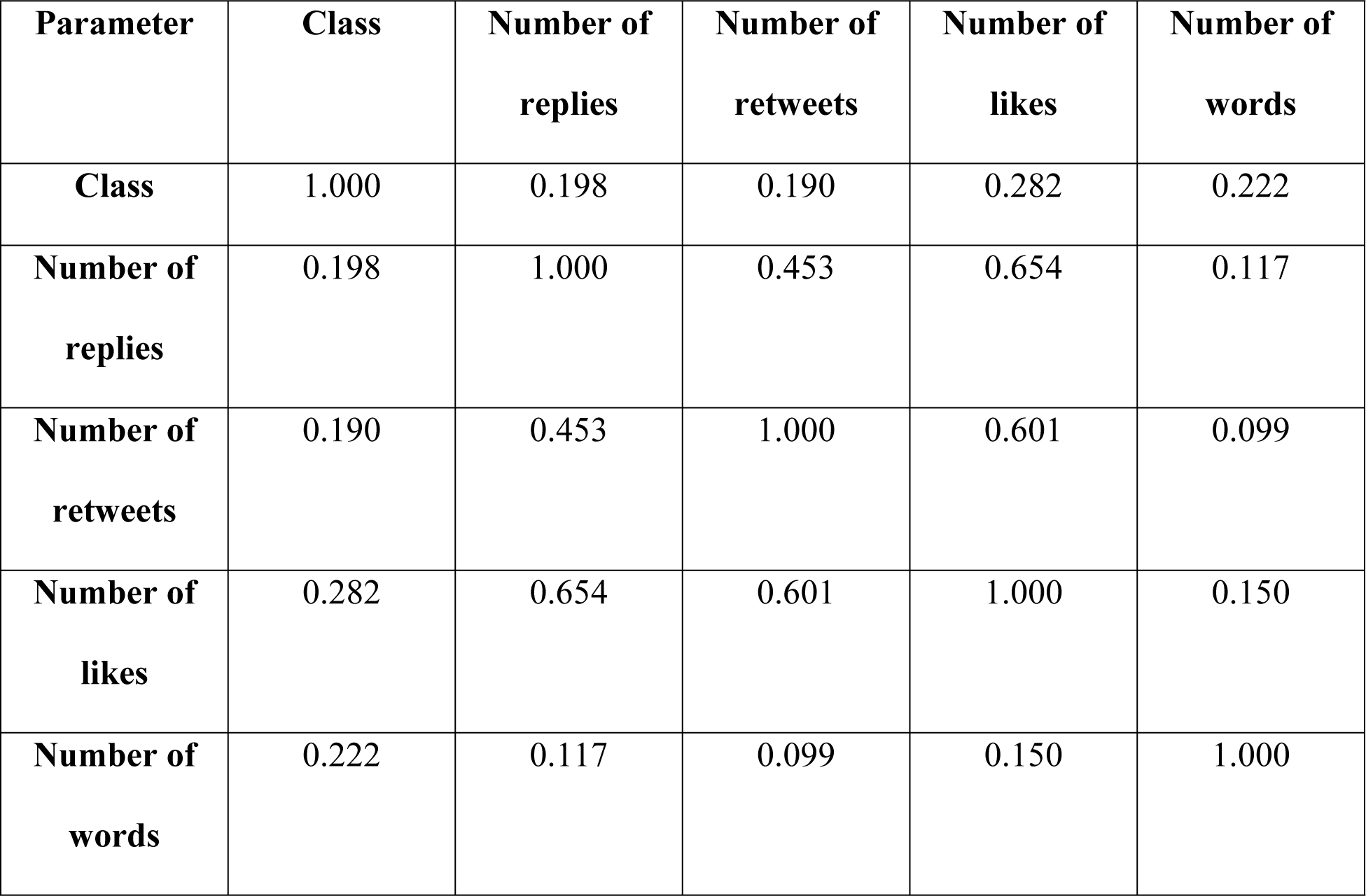
Correlation analysis results (Spearman’s correlation coefficient)

### 3.4. The validation of the machine learning algorithms

Performance metrics obtained after validating the 6 predictive algorithms (SVM, MLP, RF, the ensemble model – SVM + MLP, RCNN and BERT) are shown in Table 4. All 3 validation types are presented – repeated cross-validation, internal period validation and external validation.

**Table 4.** Validation results for the machine learning algorithms.

| **Cross-validation** |  |  |  |  |  |  |
| --- | --- | --- | --- | --- | --- | --- |
| **Metric** | **SVM** | **MLP** | **RF** | **Ensemble**<br>**(SVM + MLP)** | **RCNN** | **BERT** |
| **Classification validation** |  |  |  |  |  |  |
| Accuracy | 0.657 | 0.603 | 0.595 | 0.614 | 0.623 | 0.689 |
| Precision | 0.641 | 0.587 | 0.600 | 0.598 | 0.609 | 0.693 |
| Recall | 0.634 | 0.588 | 0.593 | 0.599 | 0.602 | 0.667 |
| F1 Score | 0.632 | 0.585 | 0.583 | 0.596 | 0.599 | 0.658 |
| Matthews Correlation<br>Coefficient | 0.480 | 0.400 | 0.401 | 0.416 | 0.425 | 0.535 |
| **Probability prediction validation** |  |  |  |  |  |  |
| ROC AUC Score (OVO) | 0.813 | 0.782 | 0.779 | 0.797 | 0.770 | 0.849 |
| ROC AUC Score (OVR) | 0.825 | 0.788 | 0.783 | 0.803 | 0.779 | 0.858 |
| **Internal period validation** |  |  |  |  |  |  |
| **Metric** | **SVM** | **MLP** | **RF** | **Ensemble**<br>**(SVM + MLP)** | **RCNN** | **BERT** |
| **Classification validation** |  |  |  |  |  |  |
| Accuracy | 0.567 | 0.546 | 0.551 | 0.576 | 0.537 | 0.601 |
| Precision | 0.572 | 0.539 | 0.554 | 0.562 | 0.525 | 0.632 |
| Recall | 0.552 | 0.536 | 0.526 | 0.557 | 0.519 | 0.573 |
| F1 Score | 0.532 | 0.520 | 0.515 | 0.542 | 0.512 | 0.539 |
| Matthews Correlation Coefficient | 0.352 | 0.317 | 0.308 | 0.354 | 0.291 | 0.416 |
| **Probability prediction validation** |  |  |  |  |  |  |
| ROC AUC Score (OVO) | 0.744 | 0.738 | 0.727 | 0.745 | 0.702 | 0.787 |
| ROC AUC Score (OVR) | 0.756 | 0.743 | 0.732 | 0.754 | 0.710 | 0.797 |
| **External validation** |  |  |  |  |  |  |
| **Metric** | **SVM** | **MLP** | **RF** | **Ensemble**<br>**(SVM + MLP)** | **RCNN** | **BERT** |
| **Classification validation** |  |  |  |  |  |  |
| Accuracy | 0.680 | 0.648 | 0.490 | 0.688 | 0.480 | 0.670 |
| Precision | 0.661 | 0.581 | 0.480 | 0.636 | 0.529 | 0.622 |
| Recall | 0.691 | 0.595 | 0.465 | 0.649 | 0.499 | 0.630 |
| F1 Score | 0.655 | 0.583 | 0.456 | 0.638 | 0.470 | 0.606 |
| Matthews Correlation Coefficient | 0.530 | 0.435 | 0.231 | 0.504 | 0.271 | 0.490 |
| **Probability prediction validation** |  |  |  |  |  |  |
| ROC AUC Score (OVO) | 0.818 | 0.772 | 0.736 | 0.800 | 0.718 | 0.806 |
| ROC AUC Score (OVR) | 0.843 | 0.796 | 0.756 | 0.826 | 0.727 | 0.829 |

The distribution of predicted probabilities generated with all 6 predictive models for the external dataset is illustrated in Figure 3.

**Fig 3.**
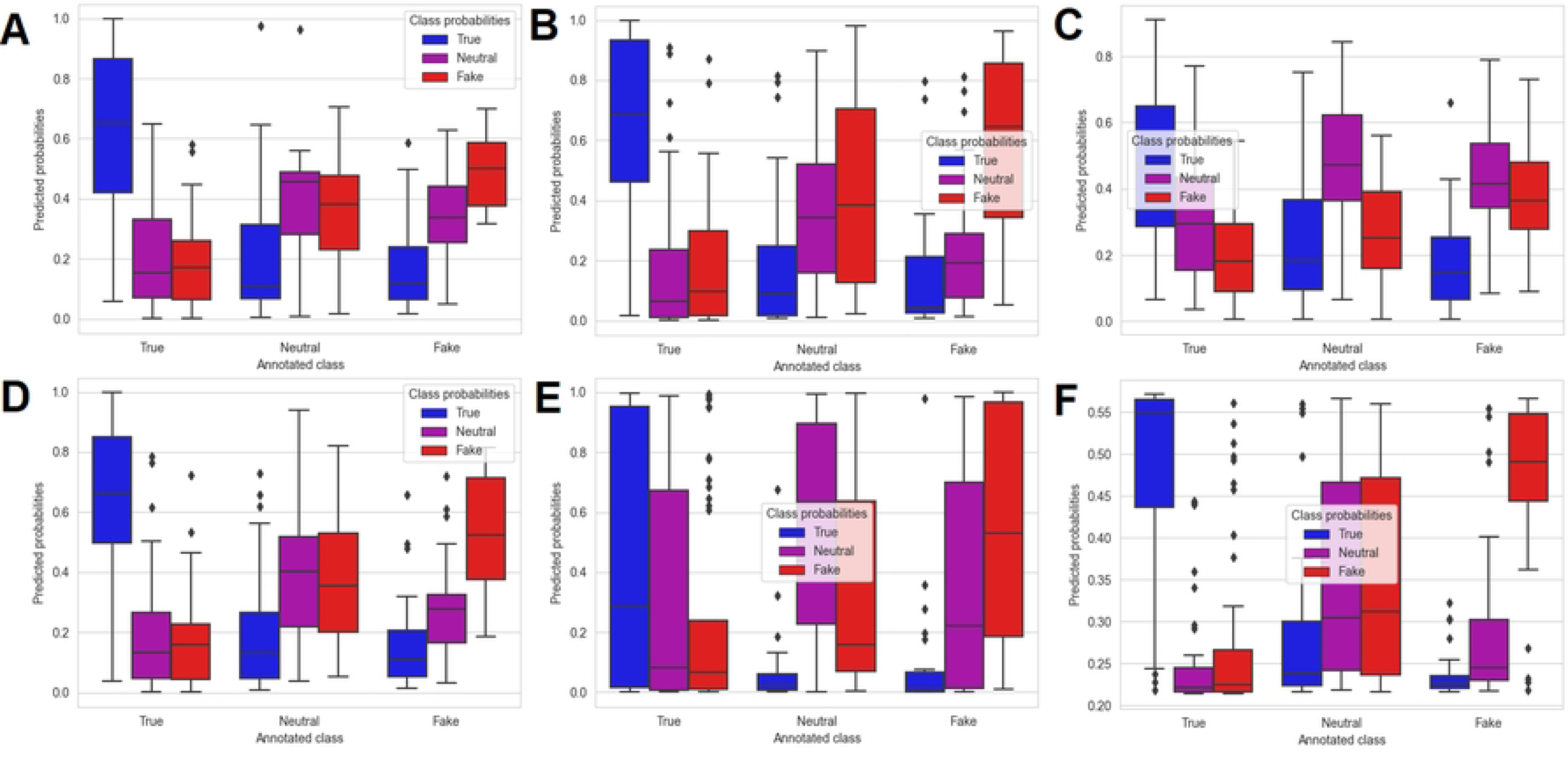
Boxplot representation of the predicted probabilities for the machine learning algorithms obtained on the external dataset: SVM algorithm (A), MLP algorithm (B), RF algorithm (C), Ensemble algorithm (SVM + MLP) (D), RCNN algorithm (E), BERT algorithm (F)

### 3.5. Implementation example of the SVM algorithm

The final algorithm chosen for implementation was the Support Vector Classifier, due to its enhanced predictions quantified through the ROC AUC Score (Table 4). Table 5 presents the probabilities returned by the algorithm for 3 tweets from the external dataset (one true tweet, one neutral tweet and one fake tweet). In order to comply with the General Data Protection Regulation, the exemplified tweets were translated and partially reformulated. In addition, in order to allow a better understanding and exemplification of tweet structure and machine learning predictive abilities, the probabilities for nine extra tweets are presented in supplementary S1 Table (three true tweets, three neutral tweets and three fake tweets).

**Table 5.** Detailed example of implementation of SVM algorithm on 3 tweets from the external dataset.

| **Reformulated tweet content** | **Predicted probability (true)** | **Predicted probability (neutral)** | **Predicted probability (fake)** | **Predicted class** | **Annotated class** |
| --- | --- | --- | --- | --- | --- |
| **Tweet A:**<br>At 10 days after the second Covid vaccine shot, the risk of getting infected is very low. | 72.56% | 6.79% | 20.66% | Class 0<br>(true) | Class 0<br>(true) |
| **Tweet B:**<br>I got the vaccine, mind your own business. | 9.94% | 43.38% | 46.68% | Class 2<br>(fake) –<br>erroneous<br>prediction | Class 1<br>(neutral) |
| **Tweet C:**<br>Mass vaccination would be catastrophic for humankind. | 11.60% | 23.02% | 65.37% | Class 2<br>(fake) | Class 2<br>(fake) |

The validation of the machine learning predictive algorithms yielded modest results in terms of classification evaluation (Table 4). The Matthews Correlation Coefficient, considered the most important metric used to assess the discriminative power of the implemented models, yielded values ranging from 0.4 to 0.535 for the cross-validation technique, from 0.308 to 0.416 for the internal period validation, as well as from 0.231 to 0.53 for the external validation of the developed algorithms. Overall, by averaging the 2 types of internal validation, BERT resulted in the highest Matthews Correlation Coefficient, of 0.535 for the cross-validation and 0.416 for the internal period validation, with a 5.5% increase for cross-validation, as well as a 6.2% increase for internal period validation as compared to SVM (which yielded a 0.48 Matthews Correlation Coefficient for the cross-validation and a 0.416 value for the internal period validation). However, it should be noted that on the external validation, the SVM algorithm outperformed BERT in terms of raw classification ability, with a 0.53 Matthews Correlation Coefficient, while BERT yielded a value of 0.49 for this validation metric. Since the Matthews Correlation Coefficient is a particular case of the Pearson product-moment correlation coefficient, its values have the same interpretation and hence it can be stated that in most cases, the validation of the BERT and SVM algorithms (the best models with regards to the internal and external validation respectively) yielded moderate to moderately high positive correlations between the true and predicted labels [35].

Nevertheless, the most important evaluation of the machine learning models was represented by the probability prediction evaluation, which tested the ability of the algorithms of estimating well calibrated probabilities, as quantified though the ROC AUC Score (using both an OVO and an OVR approach). As for the Matthews Correlation Coefficient, the ROC AUC Score yielded the lowest results for the internal period validation (OVO ROC AUC Score ranged from 0.702 to 0.787; OVR ROC AUC Score ranged from 0.71 to 0.797), followed by the external validation (OVO ROC AUC Score ranged from 0.718 to 0.818; OVR ROC AUC Score ranged from 0.727 to 0.843), while the cross-validation resulted in the highest ROC AUC values (OVO ROC AUC Score ranged from 0.77 to 0.849; OVR ROC AUC Score ranged from 0.779 to 0.858). Similar to the results obtained in terms of Matthews Correlation Coefficient, the highest ROC AUC Scores were obtained in case of BERT for both internal validation strategies (for cross-validation: OVO ROC AUC = 0.849, OVR ROC AUC = 0.858; for internal period validation: OVO ROC AUC = 0.787, OVR ROC AUC = 0.797), followed by the SVM algorithm. On the other hand, similar to the raw classification validation, SVM resulted in improved results for the ROC AUC Scores when taking into consideration the external validation (OVO ROC AUC = 0.818, OVR ROC AUC = 0.843, as opposed to a 0.806 value in case of OVO ROC AUC and a 0.829 OVR ROC AUC for the BERT model). The enhanced results which were obtained for internal validation in case of BERT might be explained that the current study implemented a pre-trained BERT model based on a large Romanian text corpus of 15 GB. Nevertheless, BERT validation yielded less accurate results than the SVM when taking the external tweets dataset into consideration, which might have been caused by a moderate amount of overfitting on the internal data (1300 tweets), as well as by the low level of complexity of the processed tweets. In addition, the RCNN model implemented through the Tensorflow library provided poor results both in terms of raw classification and probability estimation (0.702-0.710 ROC AUC Scores for internal period validation and 0.718-0.727 for external validation), which were in most cases the lowest of all 6 implemented machine learning models. These results were obtained despite the high complexity of RCNN and its ability to memorize both temporal and spatial relationships from texts. One reason for the poor results might be related to the relatively short posts which are usually distributed through the Twitter platform and to the fact that the RCNN, in contrast to the implemented BERT model, lacked a specific Romanian based text corpus and didn’t include any pretrained algorithm. Moreover, we argue that a complex model architecture (with both recurrent and convolutional layers), without any predefined recommendations, is difficult to model so that it reaches optimal results on a text corpus which contains posts in a narrowly spoken language, such as Romanian [32,33].

In terms of analysis of predicted probabilities (for the external dataset) quantified through the boxplot representations (Figure 3), both SVM (Figure 3(A)) and BERT (Figure 3(F)) offered good discrimination when comparing the estimated probabilities with the true (annotated) class. However, the main difference in the performance of the two models can be seen in the probability estimation for the tweets labelled as neutral. More specifically, the SVM offered a more accurate discrimination when predicting the probabilities that the neutral tweets from the external dataset are true, neutral or fake, the probability of being neutral being higher on average than the probability that the tweet was true or fake, which was also reflected in the lower ROC AUC Scores for BERT, when compared to SVM. By contrast, the BERT model returned on average a higher probability that the neutral tweets are fake, as compared to neutral. However, the BERT model discriminated more accurately between the true tweets, as well as the fake tweets and the rest (neutral/fake and true/neutral, respectively), while the SVM offered a more close, but still valid discrimination.

## 4. Discussion

A detailed analysis of a batch of relevant vaccine tweets from several periods within the Covid-19 pandemic was undertaken.

The preliminary analysis implied the manual annotation of a total number of 1400 tweets, as well as a preliminary analysis for establishing specific word patterns within the posts and the correlations between the manual annotation and other tweet characteristics. The supervised analysis consisted of building and validating several machine learning prediction models based on their ability of estimating the probabilities that a specific Twitter post related to vaccines is true, neutral or fake.

The manual annotation of the collected Twitter posts yielded good results in terms of inter-agreement evaluation based on Krippendorff’s alpha [38]. The inter-agreement was better for the external dataset (100 tweets, Krippendorff = 0.7) than for the internal one (1300 tweets, average Krippendorff = 0.64), partly due to the fact that the tweets from the external dataset were annotated by all 9 annotators. Moreover, the Krippendorff obtained for each of the 3 subsets within the internal data showed a certain degree of variability, with its values ranging from 0.58 to 0.69. Indeed, as with other social media posts, the ones from Twitter, even when relating to health issues, are written in a free, subjective manner, since they are mostly written by individual persons which are granted the freedom of expression [39]. Therefore, there is a high probability that the annotators ran into several ambiguous tweets and hence the interpretation of such content could have been made different depending on the content and the annotator’s subjective interpretation.

In addition, the subjective and diverse ways in which the vaccines posts were written are emphasized in Table 2, where the most relevant word patterns within the 3 classes (true, neutral and fake) are given. Interestingly, the true posts contained most often different forms of the noun „vaccine” and the verb „to vaccinate”, which could be explained by the fact that the true posts, when compared to the neutral and fake ones, contained the most references to news articles and to official data related to the Romanian vaccination campaign, such as the number of persons which were partially and fully vaccinated within a specific time period, the number of administered vaccine doses or the updated vaccine supply. In contrast, the tweets which were labelled as fake (false or manipulative information) also contained many references to the various forms of „vaccine” and „to vaccinate”, but quite often they were referenced in a subjective and manipulative manner. As an example, the word combination „experimental vaccine” was identified as one of the most relevant patterns of the fake tweets, suggesting that the Covid vaccines were not tested enough before being administered in the general population, information which is false, according to various health authorities and fact check websites. One of the main reasons of propagating such misinformation would be to make the population believe that the vaccines are dangerous for health and cause many severe adverse reactions [23,24]. In addition, words with aggressive connotation were more frequent within the tweets labelled as fake, as compared to the true and neutral ones.

In terms of correlation analysis results, the majority of obtained Spearman’s coefficients showed moderate, but statistically significant correlations (Table 3, Figure 2(D)). The manual classification (considered as class 0 – true, class 1 - neutral, class 2 – fake) was positively correlated with all of the three tweet characteristics: number of replies (r = 0.198), number of retweets (r = 0.190) and number of likes (r = 0.282). These results, even though suggesting a modest positive correlation, imply that the fake vaccine tweets have a higher impact on social media, tending to be retweeted and liked more often than the true and neutral ones (this might in turn prioritize vaccine false information even more because of the Twitter algorithm) [40]. The results are similar to the ones reflected in other studies by analyzing the online spread of misinformation [41–43]. For example, the work conducted by Vosoughi et al found among 126000 stories related to various topics that the ones labelled as false misinformation had a more pronounced spread on Twitter as compared to the valid ones [42]. Even though the effect was more pronounced for information about politics, the study raises important awareness, especially considering the fact that several studies show that the online spread of health misinformation may be, at least partially, politically driven [43]. These observations, combined with the fact that online anti-vaccine groups and accounts are more strongly connected and more likely to influence those with neutral views, make enforcing policies on limiting the spread of vaccine misinformation of an utmost importance [5,6].

Based on the OVO and OVR ROC AUC Score values which were computed for the external validation of the machine learning models (Table 4, Figure 3), the SVM algorithm was chosen for building the final predictive model [34]. In addition, all ROC AUC Scores which were obtained when validating the SVM algorithm were above 0.74, which proved the well calibrated probabilities returned by the model. Hence, from a practical point of view, the final SVM model could be used for future identification of the most relevant vaccine related Twitter posts, by sorting the automatically collected large tweet lists based on the predicted probabilities that the specific posts represent true, neutral or fake content. The obtained information, after manually analyzed, if presented through a web platform, could further aid in raising awareness regarding valid information, fake news content, as well as irrelevant information related to vaccines and shared through Twitter platform [20,34].With regards to the machine learning validation results obtained in other studies, a relevant comparison with the ones from the current studies would be difficult, since the majority of the studies which used vaccine related Twitter content reported the F1 Score as the most important classification evaluation metric [12–17,36,44,45]. The F1 Score was computed in the current study as well and can be regarded as an acceptable balanced measure between precision and recall. However, as highlighted by Chicco and Jurman, F1 Score can provide overoptimistic results when evaluating the performance of a predictive model [36]. That is the main reason for which the focus in the current study, when evaluating the classification performance of the 6 algorithms, was put on the Matthews Correlation Coefficient [35,36]. Moreover, the reported F1 Scores showed a high degree of variability, some studies reported F1 Scores of under 0.6, while others reported enhanced values, of 0.7-0.8 and others obtained almost perfect values, of over 0.95, while the implemented machine learning algorithms included both classical (such as Random Forest and SVM) and newer model types (such as deep learning and BERT) applied on various languages, such as English, French, Dutch or Moroccan [12–17,44,45]. As a comparison, the maximum F1 Score which was obtained in the current study SVM ranged from 0.542 (internal period validation – SVM+MLP Ensemble) to 0.658 (cross-validation – BERT) and 0.655 (external validation - SVM). The obtained F1 Score is therefore smaller than that reported by most of the studies; however, as was already mentioned, the most important validation metric in our study, the ROC AUC Score, yielded maximum values of over 0.8, which translates into well calibrated probabilities [34].

Another relevant example is a study which implemented an algorithm based on recurrent convolutional neural networks, with BERT as a word embedding model [46]. Even though it did not use the F1 Score as evaluation metric, it achieved superior accuracy, of 0.989, when tested on a real-world dataset, which contains real and fake news propagated during the US General Presidential Election from 2016, with over 20000 instances, both Twitter and Facebook being widely used for disinformation purpose [46,47]. Other research which aimed at detecting social media non-vaccine related disinformation implemented a hybrid deep learning model (based on recurrent neural networks) and achieved a F1 Score of 0.894, lower than in the study which specifically used BERT [46,48]. As a comparison, in our study, we obtained an accuracy of 0.601-0.689 when evaluating the BERT model; however, our dataset was much smaller and was specifically related to vaccine information distributed through Twitter.

In terms of studies which performed unsupervised analysis on specific disinformation propagated through Twitter, the research performed by Kobayashi et al is worth mentioning. It included 100 million vaccine related Japanese tweets, on which a topic, as well as a time series analysis were performed [49]. In addition, with respect to other studies which specifically evaluated social media disinformation, a study, conducted by De Clerck analyzed the general spread of disinformation through Twitter platform by taking into consideration numerous countries included in the Twitter information operations report. It proposed maximum entropy networks for identifying and quantifying specific patterns in the interactions between numerous Twitter users which might have had an important impact on spread of disinformation (whether or not health related). The analysis had the advantage of applying various algorithms and including a large number of tweets from different countries (e.g. Armenia, China, Russia, Serbia, Turkey) [50]. While our study did not implement any form of unsupervised analysis, we argue that the wordcloud representation and correlation analysis which were undertaken give context to the implemented and publicly available machine learning model.

Regarding the practical implementation of the SVM model (Table 5), the given examples provide relevant insights regarding the Romanian tweets structure, as well as the predictive algorithm use case. The first tweet (Tweet A) refers to a valid scientific information – indeed, especially considering the fact that the post was written in April 2021, when the highly contagious Omicron variant and its subvariants were not circulating, two doses of either the RNA or the viral vector vaccine (the ones which were available within the Romanian Vaccination Campaign) significantly reduced the risk of symptomatic Covid-19 [24,25]. The predictive model accurately estimated a 72.56% probability that the content is true, with only 20.66% chance of being misinformation and 6.79% of being neutral. The second tweet (Tweet B) was manually labelled as neutral, being an irrelevant statement regarding someone who got the Covid vaccine. However, the SVM algorithm erroneously classified the tweet as being fake, possibly due to the fact that it was written in a slightly aggressive manner. Nonetheless, when analyzing the predicted probabilities, the model returned a 43.38% risk that the content is neutral and a 46.68% risk that the tweet refers to false information, with only 3.3% higher than the probability of containing neutral information. The third tweet given as a practical example (Tweet C) was manually labelled as false information (fake). The algorithm returned the same classification, with a 65.37% probability that the content is fake, a 23.02% probability that it is neutral and a 11.60% chance of being true. The content of the tweet is a classical conspiracy theory, which tries to suggest that mass vaccination is not only unnecessary, but detrimental. The information is obviously false: the essential role of vaccines in leading to herd immunity and controlling infectious diseases is well established [24].

The current study has a few important advantages. First of all, to our knowledge, this is the first study analyzing vaccine fake news written in Romanian from social media posts. While Romanian is a narrowly spoken language, limited to Romania and Republic of Moldova, we argue that by providing the detailed Python code which includes the specified analyses and the developed predictive machine learning algorithm, as well as the processed (annotated, vectorized and anonymized) internal and external data, our work could be used by other researchers in future studies, with easy translation to other languages [21,51].

Secondly, as a difference from other similar studies, which used two classes (such as general information and misinformation) during the data labelling process, the current work used for manual annotation three classes (true, neutral and fake) [12,13,15–17]. It can be argued that this approach enhances the complexity of machine learning models and provides context to the social media analysis. In addition, besides the raw classification, the machine learning models which were developed provide probability estimates, a relevant feature which may aid in future selection of relevant vaccine tweets based on approaches which imply sorting the predicted probabilities, such as the ones presented in Table 5 and Supplementary Table 1. The predictive algorithms were validated in a consistent manner, both for classification and probability estimation. Relevant validation strategies were implemented: the internal period validation ensured the internal consistency of the models with regards to performing on tweets from different pandemic periods, while the external validation ensured the evaluation of the algorithms on unseen data (Table 4, Figure 4) [32–34].

Nevertheless, the current work has a series of limitations. First of all, the number of collected and annotated tweets (1300 – internal dataset, 100 – external dataset) can be regarded as very low when compared to other studies (therefore, the variability and complexity of the developed SVM algorithm could have been negatively impacted) [12–17,37]. For example, Kunneman et al conducted a study for measuring the stance towards vaccination (non-Covid vaccines: the messages were extracted prior to the pandemic period), based on a total number of 8259 annotated tweets written in Dutch; however, the study only achieved a Krippendorff’s alpha between 0.27 and 0.35, significantly lower than that from the current study [14]. However, Hayawi et al undertook a vaccine misinformation analysis based on 15073 annotated English tweets; the annotation process had the advantage of being further validated by health experts and also lead to very good machine learning validation metrics (0.97 precision, 0.98 recall, 0.98 F1 Score) [17]. Other studies focused on Covid-19 vaccine hesitancy; while they initially automatically collected large numbers of vaccine related tweets (for example, written in English, Turkish or French), the manual analysis of the content implied, as in our study, a small number of tweets (approximately 1000-2000) [18,19,44,45]. Therefore, it should be noted that while our study comprised indeed in a small dataset chosen for annotation, the fact that the tweets were chosen and annotated following a standardized methodology (selecting 4 relevant pandemic periods and eliminating the tweets with no retweets, as well as the fact that each Twitter post was classified by at least 3 annotators) could ensure reproducibility, especially considering the fact that the Python code for data preprocessing, wordcloud representation, correlation analysis and the development and validation of the machine learning predictive models, as well as the Tfidf vectorized dataset and the final SVM algorithm are publicly available at https://github.com/valeanuandrei/vaccine-tweets-ro-research [51].

Secondly, even though the results of the probability validation were satisfactory, the evaluation of the classification ability of the machine learning algorithms, especially for the internal period validation (a maximum Matthews Correlation Coefficient of under 0.42 and a maximum F1 Score of under 0.55), yielded modest results [35].

Therefore, the implemented natural language processing and data mining techniques, combined with the 12 practical examples of tweet classification and probability prediction, provide relevant insights regarding vaccine general information and misinformation spread through Twitter platform and written in Romanian. Future studies must aim at collecting a large number of tweets and classifying them based on a semi-supervised approach, in order to enhance the variability, complexity and predictive ability of the machine learning algorithm. After these steps are undertaken, an online platform might be developed, based on identifying new vaccine related Twitter content, to aid in raising awareness regarding the vaccine misinformation shared through social media and consequently reduce vaccine hesitancy [52,53].

## 5. Conclusions

A study aiming at analyzing and automatically classifying relevant vaccine related posts from Twitter content was undertaken. A total number of 1400 tweets from relevant pandemic periods were collected and manually classified as true information, neutral information or fake information related to vaccines. Both an unsupervised analysis (consisting of a wordcloud evaluation and a correlation analysis) and a supervised analysis (based on building several predictive machine learning algorithms – SVM, MLP, RF, an ensemble voting classifier: SVM + MLP, as well as complex deep learning models: RCNN and BERT) were implemented. The correlation analysis yielded moderate, but significant positive correlations between the tweets labelled as misinformation and the tweet engagement metrics, quantified through the number of replies, retweets and likes. The machine learning algorithms were mainly validated based on their ability of estimating the probability that a specific tweet is true, neutral or fake. The optimal results were obtained for the Support Vector Classifier, with a ROC AUC Score ranging from 0.744 to 0.843 and BERT, with a ROC AUC Score ranging from 0.787 to 0.858. Future studies must aim in enlarging the vaccine tweets database and optimizing the machine learning predictive abilities, in order to automatically identify and classify new vaccine related valid, neutral and false information distributed through Twitter platform.

## Data Availability

The Python code for data preprocessing, wordcloud representation, correlation analysis and the development and validation of the machine learning predictive models, as well as the Tfidf vectorized dataset and the final SVC algorithm are publicly available at https://github.com/valeanuandrei/vaccine-tweets-ro-research.

## Supporting information captions

**S1 Table:** Detailed example of implementation of SVM algorithm on 9 extra tweets from the external dataset

